# Estimating relative generation times and relative reproduction numbers of Omicron BA.1 and BA.2 with respect to Delta in Denmark

**DOI:** 10.1101/2022.03.02.22271767

**Authors:** Kimihito Ito, Chayada Piantham, Hiroshi Nishiura

## Abstract

The Omicron variant is the most transmissible variant of the severe acute respiratory syndrome coronavirus 2 (SARS-CoV-2) we had so far. The BA.1 and BA.2 sublineages of Omicron are circulating in Europe and it is urgent to evaluate the transmissibility of these sub-lineages. Using a mathematical model describing trajectories of variant frequencies that assumes a constant ratio in generation times and a constant ratio in effective reproduction numbers among variants, trajectories of variant frequencies in Denmark from November 22, 2021 to February 26, 2022 were analyzed. We found that the generation times of Omicron BA.1 and BA.2 are 0.60 (95%CI: 0.59–0.62) and 0.51 (95%CI: 0.50–0.52) of the length of that of Delta, respectively. We also found that the effective reproduction number of Omicron BA.1 is 1.99 (95% CI: 1.98–2.02) times and that of Omicron BA.2 is 2.51 (95% CI: 2.48– 2.55) times larger than the effective reproduction number of Delta. The generation times of Omicron BA.2 is 0.85 (95% CI:0.84–0.86) the length of that of BA.1 and that the effective reproduction number of Omicron BA.2 is 1.26 (95% CI:1.25–1.26) times larger than that of Omicron BA.1. These estimates on the ratio of generation times and the ratio of effective reproduction numbers has epidemiologically important implications. The duration of quarantine for people who contacted with an Omicron BA.1 and BA.2 patient can be reduced to 60% and 51% of that for Delta, respectively. The control measures against Omicron BA.1 and BA.2 need to reduce contacts between infectious and susceptible people respectively by 50% (95% CI: 49–50%) and 60% (95% CI: 60–61%) compared to that against Delta to achieve the same effect on their control.

## Introduction

The Omicron variant is the most transmissible variant of the severe acute respiratory syndrome coronavirus 2 (SARS-CoV-2) we had so far. The variant was designated as a variant of concern by the World Health Organization (WHO) on November 26, 2021 (World Health Organization, 2021). As of February 7, 2022, infections by Omicron were reported to WHO from official sources of 159 countries (World Health Organization, 2022). The rapid replacement of Delta by Omicron followed by a steep rise in SARS-CoV-2 infections has been observed after the introduction of Omicron in many countries, indicating that Omicron has considerably higher transmissibility than Delta. Furthermore, the BA.2 sublineage of Omicron is replacing the BA.1 sublineage in Denmark. Although attenuated disease severity of Omicron was reported (Halfmann et al., 2022; Shuai et al., 2022), there is an urgent need to evaluate the transmissibility of these Omicron sub-lineages.

Transmissions of a new variant should be characterized by two factors associated with the transmission of the virus. The first factor is effective reproduction number (*R*_*t*_), which measures how many secondary cases are generated by a single primary case at time *t*. The second factor is the generation time distribution *f*(*τ*), which describes how secondary infections are distributed as a function of time *τ* since the infection of primary case (Fraser, 2007).

Alex Selby has estimated the generation time of Omicron using case counts in England in 2021 (Selby, 2022). In a logistic regression assuming that the generation time of Omicron is the same of that of Delta, the log odds of Omicron and Delta frequencies should be on a straight-line relationship. However, the case counts in England showed a bend in the log odds of observations that deviated from its theoretical straight line. Using this bend, Selby estimated the ratio of the mean generation time of Omicron to the mean generation time of Delta to be 0.46 (95% confidence intervals (CI): 0.38–0.61).

Laung et al. have developed a mathematical model describing trajectories of variant frequencies that assumes constant ratios of effective reproduction numbers among variants (Leung, Lipsitch, Yuen, & Wu, 2017; Leung, Shum, Leung, Lam, & Wu, 2021). This method needs daily numbers of infections of each variant. In our previous paper we have proposed an approximated form of the model which is free from using daily numbers of infections (Kimihito Ito, Chayada Piantham, & Hiroshi Nishiura, 2021). Using the model, we have analyzed nucleotide sequences of SARS-CoV-2 collected in Denmark (K. Ito, C. Piantham, & H. Nishiura, 2021). This model, however, assumed that generation times of variants followed the same probability distribution and that effective reproduction numbers of Omicron BA.1 and BA.2 were the same. In this paper, we propose a new model in which the generation times of variants may be different from each other. Analyzing latest variant count data observed in Denmark using the new model, we compare generation times and effective reproduction numbers of Omicron with those of Delta and compare effective reproduction numbers of Omicron BA.1 and Omicron BA.2 with respect to (w.r.t.) Delta.

## Materials and Methods

### Model of variant frequencies

Consider a situation where viruses of variant *a* are circulating in the population at the beginning of the target period of analysis and variant *A*_1_ and *A*_2_ are newly introduced to the same population at calendar times 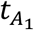 and 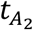, respectively. Let 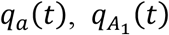, and 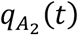 be frequencies of variant *a, A*_1_, and *A*_2_ in the viral population at calendar time *t*, respectively. Since there is no infection of *A*_1_ before its introduction, 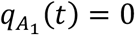 for a calendar time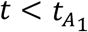, and the same is true for *A*_1_ for 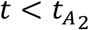.

The dynamics on the frequencies of infections by *a, A*_1_, and *A*_2_ is determined by the generation times and effective reproduction numbers of infections of the two variants. We assume that the generation time (GT) of infections by *A*_1_ and *A*_1_ are *c*_1_ and *c*_2_ times longer than that of *a*, respectively. We call the value of *c*_*i*_ the relative generation time of *A*_*i*_ with respect to *a*. Let 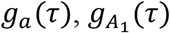, and 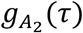 be probability density functions of generation time distributions of infections by *a, A*_1_, and *A*_2_, respectively. The assumption on generation times can be described by the following equation:

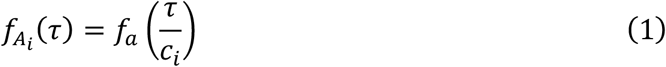

for any generation time *τ* ≥ 0. To deal with the calendar time system, we truncate the generation time distributions at *τ* = 1 and *τ* = *l* and discretize it as follows:

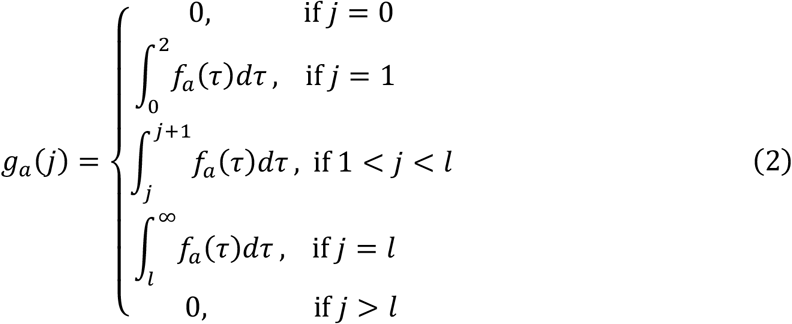

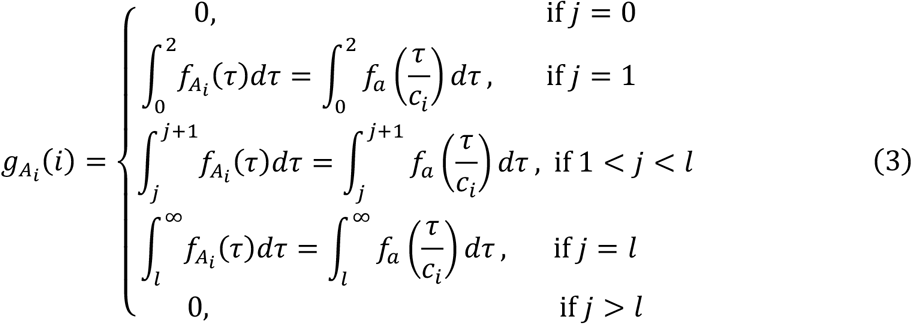

Let *I*(*t*) be the total number of new infections by either *a, A*_1_, or *A*_2_ at calendar time *t*, and let *R*_*a*_(*t*), *R*_*A*_(*t*), and *R*_*A*_ (*t*) be effective reproduction numbers of variants *a, A*_1_, and *A*_2_ at calendar time *t*, respectively. From the definition of the instantaneous reproduction number (Fraser, 2007), 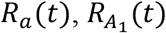, and 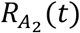 can be written as follows:

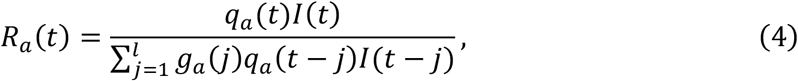

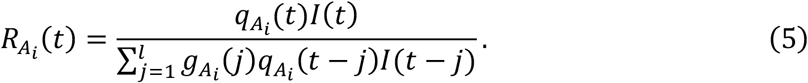

We assume that a patient infected by *A*_1_ and *A*_2_ generates respectively *k*_1_ and *k*_2_ times as many secondary transmissions as those of a patient infected by *a* regardless of *t*. Using effective reproduction numbers of *a, A*_1_, and *A*_2_, this assumption can be described by the following equation:

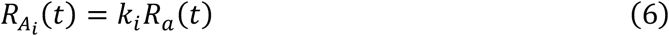

for each calendar time 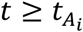. The value of the constant *k*_*i*_ is called relative reproduction number of *A*_*i*_ w.r.t. *a*.

Under the two epidemiological assumptions described by Equations (1) and (6), the frequency of variant *A*_*i*_ in the viral population at time *t* is now modeled as

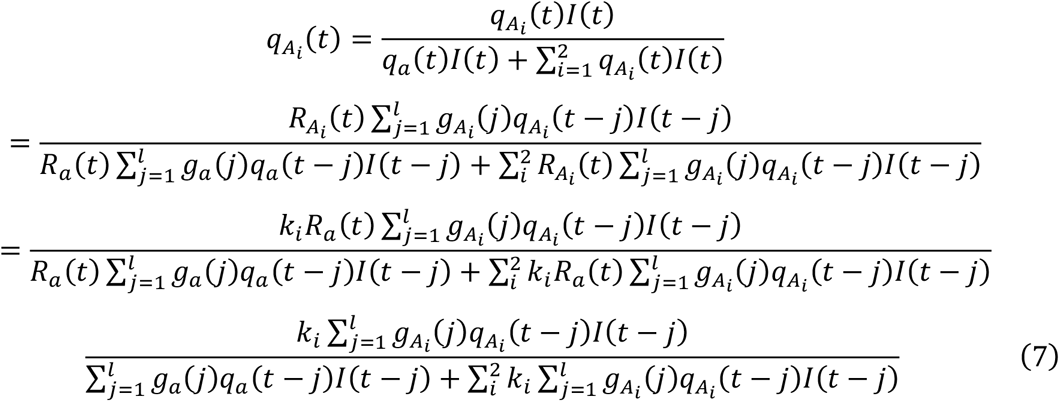

To allow statistical estimation of *c*_*i*_ and *k*_*i*_ without knowing *I*(*t*), we assume that the number of new infections does not greatly vary within a single generation of transmission from time *t* − *l* to *t* − 1, i.e.

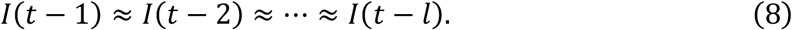

for each calendar time *t*. Then we obtain our main formula:

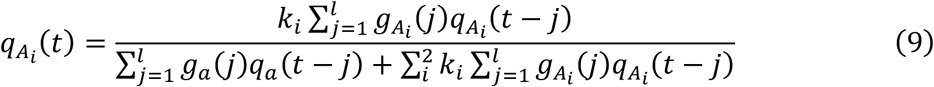

Note that Equation (8) is used just for approximating Equation (7) with Equation (9) for each calendar time *t*. We do not assume that *I*(*t*) is constant during the entire period of the analysis, although a chain of the assumptions leads such a consequence.

### Model of observation counts

Let 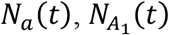, and 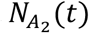 be the number of variants *a, A*_1_, and *A*_2_ observed at calendar time *t*. Assuming that variants *a, A*_1_, and *A*_2_ is sampled following a multinomial distribution with probabilities 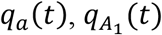, and 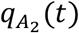 in 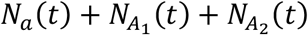 trials, parameters *c*_*i*_ and *k*_*i*_ and the initial variant frequencies of 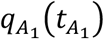 and 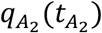, can be estimated by maximizing the likelihood function of the multinomial distributions, which is described as follows:

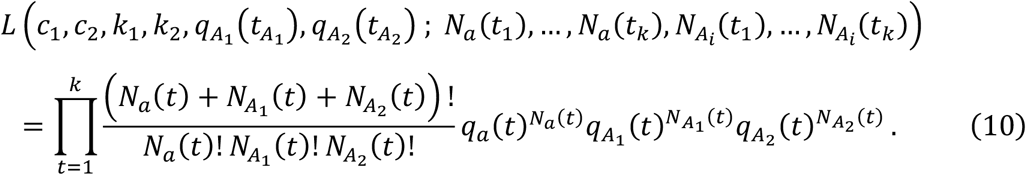

### Observation of variants in Denmark

As of February 28, 2022, the earliest Danish Omicron sequences registered in the GISAID database (Shu & McCauley, 2017) were sampled on November 22, 2021. We downloaded a total of 150,164 nucleotide sequences of SARS-CoV-2 viruses sampled in Denmark during November 22, 2021 to February 22, 2022 were downloaded from the GISAID database on February 28, 2022. Of these, 47,698 were labeled as PANGO lineages corresponding to Delta (Rambaut et al., 2020) and 102,222 were labeled as lineages corresponding to Omicron. Of sequences corresponding to Omicron, 36,118 were labeled as BA.1, 66,076 were BA.2, seven were BA.3, and 21 were B.1.1.529. The other 244 sequences were labeled as lineages corresponding to neither Delta nor Omicron. The 272 sequences that were not labeled as Delta, BA.1 or BA.2 were ignored in the subsequent analyses. We consider Delta as the baseline variant *a* and Omicron BA.1 and BA.2 as *A*_1_ and *A*_2_ in our model. We set 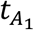 And 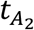 to November 22, 2021 and December 5, 2021 which are the sample collection date of the earliest Omicron BA.1 and BA.2, respectively. For each date *t* from November 22, 2021 to February 22, 2022, the number of sequences labeled as Delta collected at that day was used as *N*_*a*_(*t*), that of Omicron BA.1 as 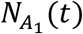 and that of Omicron BA.2 as 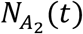. Dates of sample collection and PANGO lineages assigned to sequences are provided in Supplementary Table 1.

### Generation times of the Delta variant

Hart et al. reported that the distribution of the generation time of the Delta variant had a mean of 4.7 days with a standard deviation of 3.3 days by analyzing household transmission data from UK Health Security Agency (Hart et al., 2021). Based on the mean and the standard deviation, the generation times of infections by Delta were assumed to follow the gamma distribution with a shape parameter of 2.03 and a scale parameter of 2.32. We used the probability density function of this gamma distribution as *f*_*a*_(*τ*).

### Maximum likelihood estimation of parameters

We estimated the parameters of our model from the dataset using three assumptions: The One-GT model, the Two-GT model, and the Three-GT model. The Three-GT model assumes that the GT distribution of Omicron BA.1 and BA.2 may be different from Delta in a manner described by Equation (1) and that the reproduction number of Omicron BA.1 and BA.2 may be different in a manner described by Equation (6). More precisely, *c*_1_ may be different from *c*_2_ and *k*_1_ may be different from *k*_2_. The Two-GT model assumes that BA.1 and BA.2 shares the same generation time distribution. In other words, *c*_1_ must be equal to *c*_2_ and *k*_1_ may be different from *k*_2_. The One-GT model assumes that the GT distributions of Omicron BA.1 and BA.2 infections are the same as that of Delta, i.e. *c*_1_ = *c*_2_ = 1, and that the reproduction number of Omicron may be different in a manner described by Equation (6). Note that *c*_1_ = *c*_2_ in the Three-GT model implies the Two-GT model and that *c*_1_ = *c*_2_ = 1 in the Three-GT model implies the One-GT model. The 95% CIs are calculated using the profile likelihood method (Pawitan, 2013). The maximum likelihood estimation and the 95% CI calculation were done using daily count data from November 22, 2021 to February 22, 2022. We used the augmented Lagrangian algorithm implemented in the NLopt module of the Julia language (Conn, Gould, & Toint, 1991) for the maximum likelihood estimation and the calculation of 95% CIs of parameters. Models were compared using the Akaike information criterion (AIC) (Akaike, 1974).

## Results

The relative generation time of Omicron w.r.t. Delta, *c*_*i*_, and the relative reproduction number *k*_*i*_, were estimated from the daily count of Delta and Omicron BA.1 and BA.2 in Denmark (Table 1). The Three-GT model estimated *c*_1_ and *c*_2_ to be 0.60 (95%CI: 0.59–0.62) and 0.51 (95%CI: 0.50–0.52) and *k*_1_ and *k*_2_ to be 1.99 (95% CI: 1.98–2.02) and 2.51 (95% CI: 2.48– 2.55), respectively. The Two-GT model estimated *c*_1_(= *c*_2_) to be 0.63 (95%CI: 0.61–0.67), *k*_1_ and *k*_2_ to be 2.03 (95%CI: 1.99–2.06) and 2.63 (95%CI: 2.57–2.72), respectively. The One-GT model estimated *k*_1_ and *k*_2_ to be 2.40 (95%CI: 2.38–2.42) and 3.66 (95%CI: 3.63– 3.69, respectively.

**Table 1.**
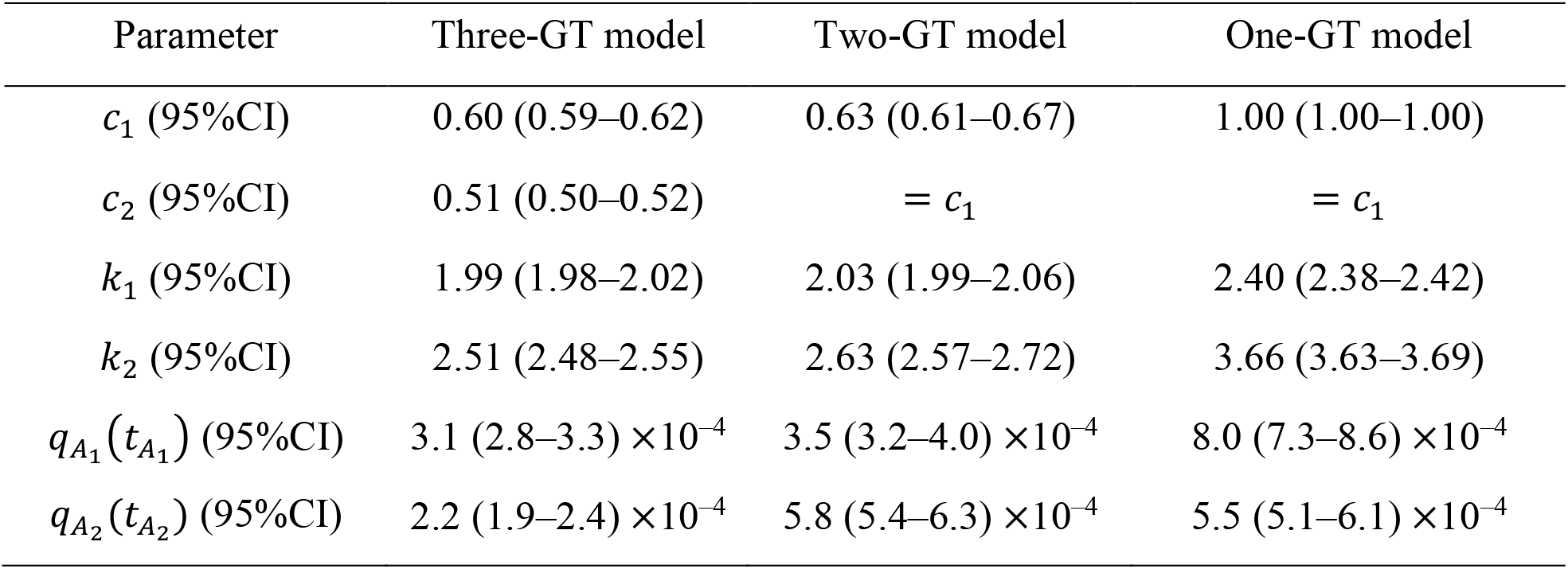
Results of parameter estimation

The maximum log likelihood of the Three-GT model was –651.4 with an AIC of 1320.8 and the Two-GT model was –670.4 with an AIC of 1356.8 while the maximum log likelihood of the One-GT model was –797.6 with an AIC of 1356.8 (Table 2). The values of AIC suggested that the selective advantage of Omicron over Delta can be decomposed into advantage in the speed of transmissions and advantage in the number of transmissions. When comparing the Three-GT model with Two-GT model, values of AIC indicate the Three-GT model was more likely than the Two-GT models. From values of *c*_1_,*c*_2_, *k*_1_, and *k*_2_ in the Three-GT model, it is suggested that the generation times of Omicron BA.2 is 0.85 (95% CI:0.84–0.86) of the length of that of BA.1 and that the effective reproduction number of Omicron BA.2 is 1.26 (95% CI:1.25–1.26) times larger than that of Omicron BA.1 under the same epidemiological condition.

**Table 2.** Akaike information criterion (AIC) of assumed models

| Model | Number of parameters | Maximum log likelihood | AIC |
| --- | --- | --- | --- |
| Three-GT | 9 | −651.4 | 1320.8 |
| Two-GT | 8 | −670.4 | 1356.8 |
| One-GT | 7 | −797.6 | 1356.8 |

Figure 1 shows trajectories of frequencies of Delta, Omicron BA.1, and BA.2 estimated using the Three-GT model and the One-GT model. The One-GT model (Figure 1(B)) shows a discrepancy between estimation and observation. At the beginning of December 2021, the One-GT model underestimated the frequency of Delta, while the overestimation can be observed from the end of December 2021 to the beginning of January 2022. On the other hand, the Three-GT model shows a good fit from the beginning to the end of the analyzed period (Figure 1 (A)). The large difference in log likelihood between the Three-GT model and One-GT model is attributed to the discrepancy between estimated frequencies and observed frequencies, and this is the reason why our model can estimate the relative generation time from trajectory of variant frequencies. The difference in the log Likelihood between the Three-GT model and Two-GT model suggested that the trajectory estimated using the Two-GT model showed overestimation and underestimation in the same way as the One-GT model, although it was difficult to visualize them.

**Figure 1.**
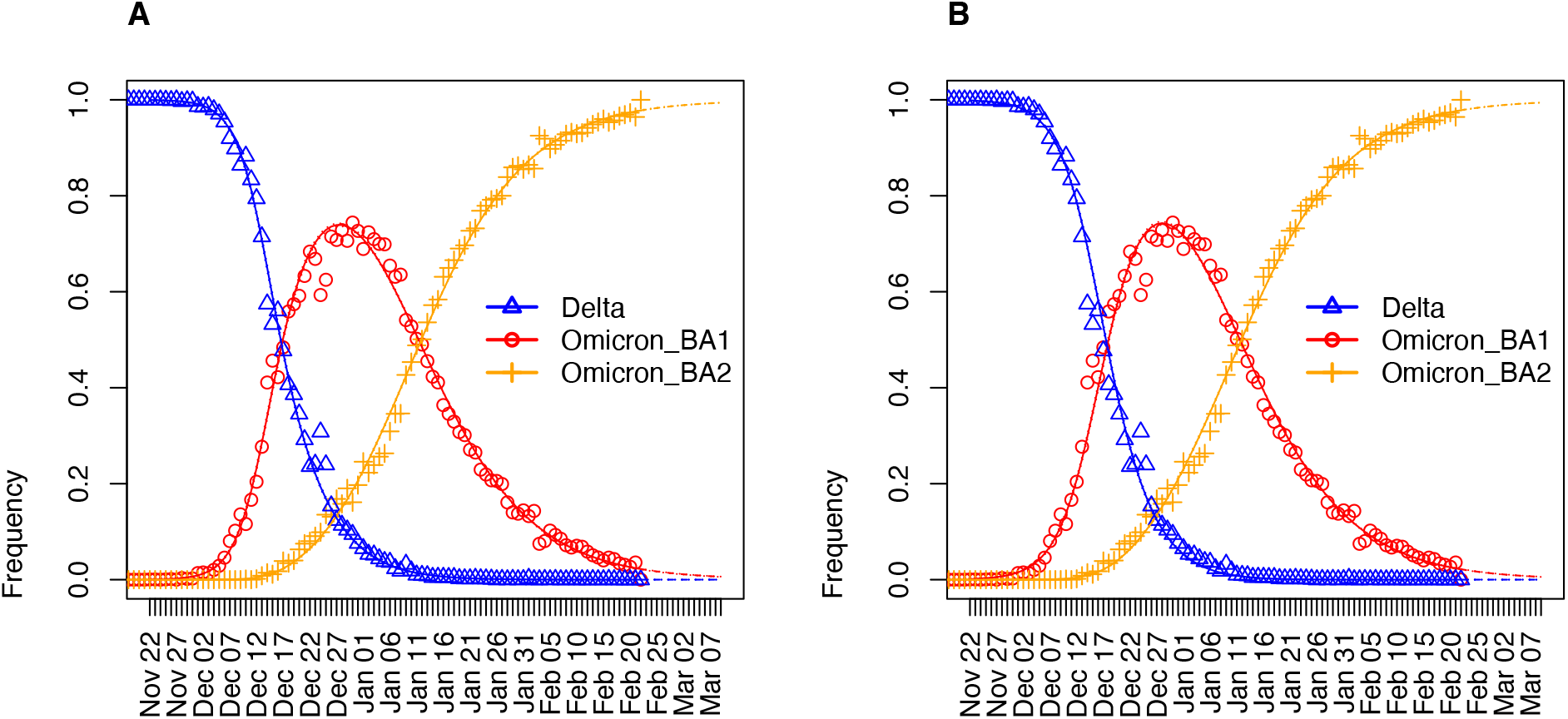
Frequencies of Delta and Omicron from November 22, 2021 to March 9, 2022 estimated using the Three-GT model (A) and the One-GT model (B). Triangles, circles, plus signs represent the frequencies of Delta, Omicron BA.1, and BA.2, respectively. Solid lines indicate the maximum likelihood estimates of frequencies of Delta (blue), Omicron BA.1 (red), and Omicron BA.2 (orange). Dashed lines from February 23, 2022 to March 9, 2022 indicate predicted frequencies of Delta (blue), Omicron BA.1 (red), and Omicron BA.2 (orange). Dotted lines show the upper bound and lower bound of 95% confidence intervals of estimated and predicted frequencies for Delta (blue), Omicron BA.1 (red), and Omicron BA.2 (orange).

Figure 2 shows the population average of the relative generation time and relative reproduction number of SARS-CoV-2 infections w.r.t Delta estimated from variant observations in Denmark using the Two-GT model. The replacement of Delta by Omicron BA.1 speeded up around December 7, 2021. From then, the population average of the generation times started decreasing and the population average of the relative reproduction numbers started increasing due to the Delta–Omicron BA.1 replacement. The decrease in the SI is saturated because when Delta was replaced by Omicron BA.1. However, population averages of relative reproduction numbers continued to increase due to the Omicron BA.1– Omicron BA.2 replacement.

**Figure 2.**
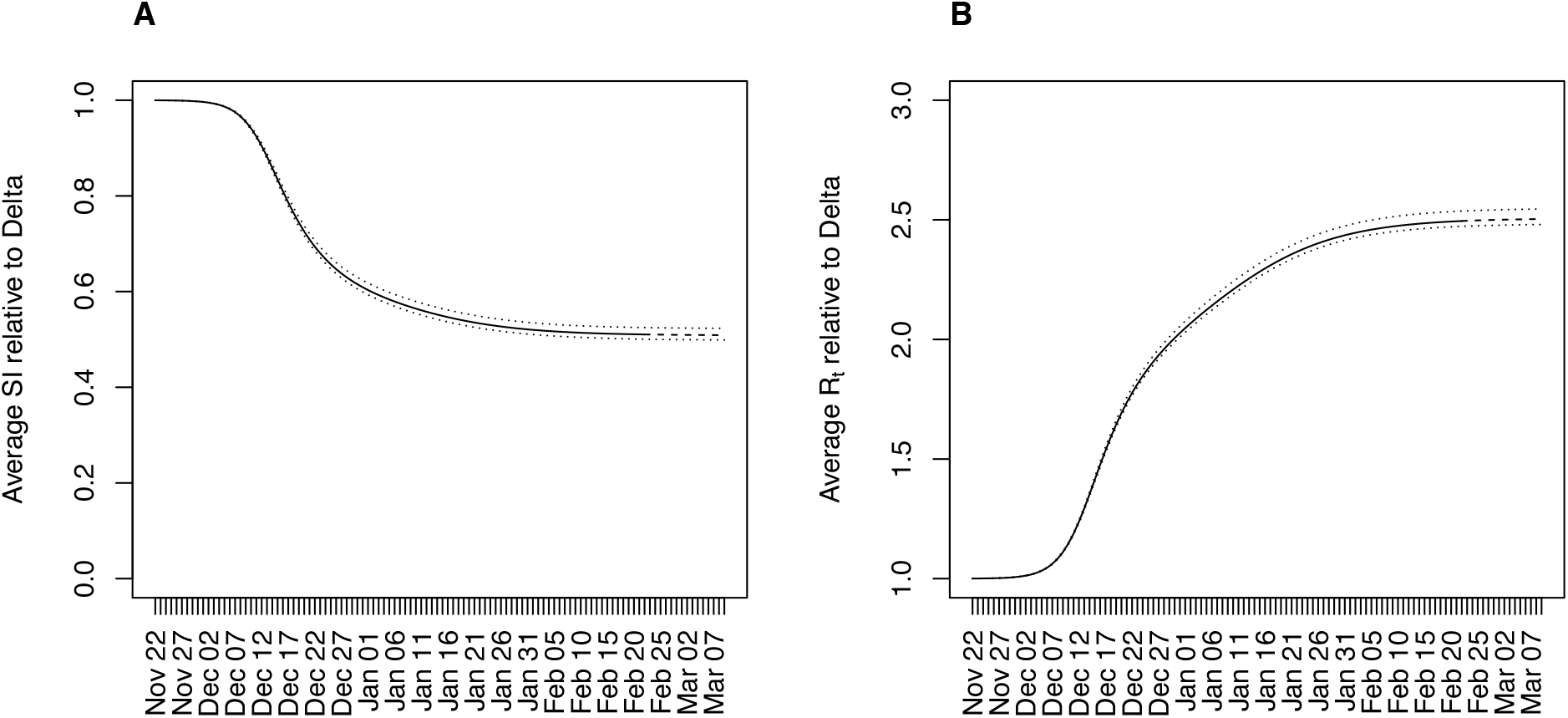
Population average of the relative generation time (A) and relative reproduction number (B) with respect to the Delta variant from November 22, 2021 to March 9, 2022. In each panel, the solid line represents maximum likelihood estimates until February 22, 2022 and the dashed line represents predicted values from February 23 to March 9, 2022. Dotted lines indicate the upper and lower bounds of the 95% confidence intervals of estimated and predicted values.

## Discussion

We have developed a mathematical model describing the time evolution of variant frequencies using both the ratio of generation times and the ratio of effective reproduction number among variants. Analyzing variant counts of SARS-CoV-2 sampled in Denmark from November 22, 2021 to December 31, 2021 using the model, we have shown that the selective advantage of Omicron BA.1 and BA.2 over Delta was decomposed into advantage in the speed of transmission and advantage in the number of transmission. The generation times of Omicron BA.1 and BA.2 are 0.60 (95%CI: 0.59–0.62) and 0.51 (95%CI: 0.50–0.52) of the length of that of Delta, respectively. The effective reproduction number of Omicron BA.1 is 1.99 (95% CI: 1.98–2.02) times and that of Omicron BA.2 is 2.51 (95% CI: 2.48– 2.55) times larger than the effective reproduction number of Delta. These estimates suggests that the generation times of Omicron BA.2 is 0.85 (95% CI:0.84–0.86) of the length of that of BA.1 and that the effective reproduction number of Omicron BA.2 is 1.26 (95% CI:1.25– 1.26) times larger than that of Omicron BA.1.

The estimation of these two parameters has important implications to the control of infections by Omicron. First, Omicron BA.1 and BA.2 respectively generates 1.99 (95% CI: 1.98–2.02) times and 2.51 (95% CI: 2.48–2.55) times more secondary transmissions than Delta under the same epidemiological conditions. Control measures against Omicron BA.1 need to reduce contacts between infectious and susceptible people by 50% (95% CI: 49–50%) compared to that against Delta to achieve the same effect on their control. In the same manner, control measures against Omicron BA.2 need to reduce contacts by 60% (95% CI: 60–61%).

Second, generation times of Omicron BA.1 and BA.2 are respectivery 0.60 (95%CI: 0.59– 0.62) and 0.51 (95%CI: 0.50–0.52) of the length of that of Delta. This suggests that the time needed for quarantine of people contacted with an Omicron patient may be reduced to 60% (95%CI: 48–53%) of time needed for people contacted with a Delta patient. The control of infections by Omicron needs to consider trade-off between the effort to prevent virus carriers to contact with others and the effort to encourage non-carrier essential workers to continue working. The estimates obtained by this study gives important evidence to establish a separated control measure for Omicron.

In our previous paper, we have estimated the relative reproduction number of Omicron w.r.t. Delta to be 3.19 (95%CI: 2.82–3.61) using variant frequencies observed in Denmark from November 1, 2021 to December 9, 2021 (K. Ito et al., 2021). The discrepancy in the relative reproduction numbers from this study is attributed to the following reason. Ito et al. 2021 assumed the same generation time for Omicron and Delta. In fact, the estimate using the One-GT model estimated relative reproduction numbers of 2.40 (2.38–2.42) for BA.1 and 3.66 (3.63–3.69) for the BA.2 w.r.t Delta. Both BA.1 and BA.2 were circulating in Denmark in December 2021, and the relative reproduction number estimated by our previous study is consistent with results of this study.

The estimations of this study completely depend on the variant counts based on the nucleotide sequences submitted to the GISAID database from Denmark. Most sequences from Demark were submitted by member laboratories of Danish Covid-19 Genome Consortium, which analyzed positive specimen from hospitals (Danish Covid-19 Genome Consortium, 2021). As of February 24, the sequencing rate from the 47th week of 2021 to the 7th week of 2022 is 7.2% (157,049 / 2,156,159). Given such a high sequencing rate, the extent of sequencing bias may be minimal. Evaluation of sequences submitted from other countries may confirm the results of present study.

## Data Availability

All data produced in the present work are contained in the manuscript.

## Acknowledgement

We gratefully acknowledge the laboratories responsible for obtaining the specimens and the laboratories where genetic sequence data were generated and shared via the GISAID Initiative, on which this research is based. The information on originating laboratories, submitting laboratories, and authors of SARS-CoV-2 sequence data can be found in Supplementary Table 1. We thank Heidi Gurung, Bradley Suchoski, and Sid Baccam in IEM in the United States of America for their help to convert our R program into the Julia language. This work was supported by the Japan Agency for Medical Research and Development (grant numbers JP20fk0108535). K.I. received funding JSPS KAKENHI (21H03490). C.P. was supported by the World-leading Innovative and Smart Education Program (1801) from the Ministry of Education, Culture, Sports, Science, and Technology, Japan. H.N. received funding from Health and Labor Sciences Research Grants (20CA2024, 20HA2007, and 21HB1002); the Japan Agency for Medical Research and Development (JP20fk0108140); JSPS KAKENHI (21H03198) and the Japan Science and Technology Agency (JST) SICORP program (JPMJSC20U3 and JPMJSC2105). The funders had no role in the study design, data collection and analysis, decision to publish, or preparation of the manuscript.

## Data Availability

Supplementary Table 1 contains all data needed to reproduce the result of this study.

## Author Contribution

KI and HI design the study. KI and CP collected data and conducted estimation. HN addressed public health implications. KI, CP, HN wrote the paper.

